# Relationships between demographic, geographic, and environmental statistics and the spread of novel coronavirus disease (COVID-19) in Italy

**DOI:** 10.1101/2020.09.18.20196980

**Authors:** Alessandro Rovetta, Lucia Castaldo

## Abstract

**Background:** Since January 2020, the COVID-19 pandemic has raged around the world, causing nearly a million deaths and hundreds of severe economic crises. In this scenario, Italy has been one of the most affected countries.

**Objective:** This study investigated significant correlations between COVID-19 cases and demographic, geographical, and environmental statistics of each Italian region from February 26 to August 12, 2020. We further investigated the link between the spread of SARS-CoV-2 and particulate matter (PM) 2.5 and 10 concentrations before the lockdown in Lombardy.

**Methods:** All demographic data were obtained from the *AdminStat Italia* website, and geographic data were from the *Il Meteo* website. The collection frequency was one week. Data on PM2.5 and PM10 average daily concentrations were collected from previously published articles. We used Pearson’s coefficients to correlate the quantities that followed a normal distribution, and Spearman’s coefficient to correlate quantities that did not follow a normal distribution.

**Results:** We found significant strong correlations between COVID-19 cases and population number in 60.0% of the regions. We also found a significant strong correlation between the spread of SARS-CoV-2 in the various regions and their latitude, and with the historical averages (last 30 years) of their minimum temperatures. We identified a significant strong correlation between the number of COVID-19 cases until August 12 and the average daily concentrations of PM2.5 in Lombardy until February 29, 2020. No significant correlation with PM10 was found in the same periods. However, we found that 40 *μg*/*m*^3^ for PM2.5 and 50 *μg*/*m*^3^ PM10 are plausible thresholds beyond which particulate pollution clearly favors the spread of SARS-CoV-2.

**Conclusion:** Since SARS-CoV-2 is correlated with historical minimum temperatures and PM10 and 2.5, health authorities are urged to monitor pollution levels and to invest in precautions for the arrival of autumn. Furthermore, we suggest creating awareness campaigns for the recirculation of air in enclosed places and to avoid exposure to the cold.

## Introduction

The Chief of the World Health Organization (WHO) has declared the COVID-19 pandemic the most severe pandemic in recent human history [1]. To date, over 200 countries have been involved, with over 29 million cases and 900,000 deaths [2]. Between February and April 2020, Italy was the most affected nation in terms of the number of new cases and deaths [3]. The severity of the emergency was so important that, despite a drastic drop in infections during the summer months, it is still among the top 20 nations afflicted by the novel coronavirus [4]. On January 23, two Chinese tourists tested positive for COVID-19 near Rome [5]. However, the patients appeared to have been readily isolated, averting extensive contagion. Toward the end of February, the situation began to accelerate and fall outside the control of the authorities. From February 21, to counter the spread of the virus, the Italian government declared various lockdowns that extended around the outbreak of Lodi, in the Lombardy region. On March 10, the lockdown went into effect nationwide [6]. For these reasons, we believe Italy is one of the main sources of information for fully understanding the behavior of SARS-CoV-2.

This is the first study to provide a complete and detailed history of the correlations between the spread of SARS-CoV-2 and demographic, geographic, and environmental statistics in Italy. From the analysis of our results, it was possible to highlight anomalous and/or local properties and behaviors of the virus as well as test the statistical significance of the hypotheses and scenarios proposed by other research.

## Methods

We collected data on Italian demographic statistics and pollution from the *AdminStat Italia* website and a previously published article [7, 8]. We looked for significant Pearson (*R*) and Spearman (*r*) correlations between COVID-19 cases per province in each region from February 26 until August 12, 2020, and birth rate, median age, population density, death rate, old age index, population number, percentage of unmarried people, family members, growth rate, percentage of divorcees, percentage of foreigners, foreign growth rate, and percentage of widowers. The frequency of data collection was one week. For each correlation identified, we calculated the angular coefficient of the interpolating line and correlated the latter with geographical characteristics such as regions’ latitude and minimum temperatures in February and March (last 30 years of historical data). We collected geographic data from the *Il Meteo* website [9]. The survey was conducted in the following regions: Abruzzo, Calabria, Campania, Emilia-Romagna, Friuli Venezia Giulia, Lazio, Liguria, Lombardia, Marche, Piemonte, Sardegna, Sicilia, Toscana, Veneto (number of provinces > 3). In the *Results* section, we report only significant results. The week in which the correlation approached the threshold of statistical significance is reported in parentheses, for example, Abruzzo (4). Finally, we have deepened the results of a previous study on the search for correlations between the daily average concentrations of particulate matter (PM) 10 and 2.5 until February 29 and COVID-19 cases for every province in the Lombardy region from February 26 to August 12, 2020 [7].

### Statistical analysis

Each result was reported together with its standard deviation (*SD*) and p-value (*p*); we chose a statistical significance threshold of *α* ≤ 05. For each sample, kurtosis (*k*) and skewness (*s*) were calculated using Microsoft Excel 2020 software. We used the formulas 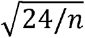 and 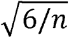, with n sample size, to obtain their respective standard deviations [10]. We considered data compatible with a normal distribution only when *t*_*k*_ = *k* · (24/*n*)^-1/2^ ≤ 1.5 and *t*_*s*_ =*s* · (6/*n*)^-1/2^ ≤ 1.5; here, the Pearson correlation index was deemed more reliable. When *t*_*k*_ ∈ [1.5, 3], *t*_*s*_ ≤ 3 or *t*_*k*_ ≤ 3, *t*_*s*_ ∈ [1.5, 3], we considered it appropriate to evaluate both correlations. Finally, when *t*_*k*_, *t*_*s*_ > 3, we judged the Spearman correlation index to be more appropriate. When we highlighted linear correlations, we used the *Igor Pro 6*.*37* software to interpolate the data through the equation *y* = *a* + *bx*. In the results section, we report the mean values *R*_*best*_, *r*_*best*_ of the correlations identified, with their 95% confidence intervals. When the coefficients exceeded the value. 700 with p ≫ .05, they were reported to specify the absence of statistical significance. To verify the importance of the correlation, we constructed a suitable correlation matrix. We have defined *pure* correlations independent from other quantities correlated with COVID-19.

## Results

### Correlation between COVID-19 cases and population number

We found significant strong correlations between COVID-19 cases and population number in 60.0% of regions such as Calabria (5), Campania (1), Lazio (1), Liguria (2), Lombardy (4), Piedmont (2), Sardinia (3), Sicily (1), and Veneto (4) (*R*_*best*_ = .935, 95% *CI*: .830 – 1.000, *p*_*best*_ = .046, 95% *CI*: .006 - .040). We identified suspicious correlations in Emilia-Romagna, Marche, and Puglia (Table 1). Since correlations have occurred in many regions from the first weeks, it is likely that the virus had spread as early as January. On the contrary, Veneto and the most affected area, Lombardy, seem to have experienced a gradual contagion. The average of the angular coefficients resulting from the linear interpolations of the pairs (COVID-19 cases, population number) is *b* = .0037 (95% *CI*. 0009, .0065).

**Table 1.** Pearson and Spearman correlations between COVID-19 cases and population density and between COVID-19 cases and population numbers.

| n | Region | $t_{k,cov}$ | $t_{s,cov}$ | Population | $R$ | $p$ | $r$ | $p$ | $t_k$ | $t_s$ |
| --- | --- | --- | --- | --- | --- | --- | --- | --- | --- | --- |
| 1 | ABRUZZO | 0.58 | 0.74 | density | .971 | .029 | .800 | .200 | 0.59 | 0.18 |
|  |  |  |  | number | .212 | .788 | .800 | .200 | 1.29 | 1.43 |
| 2 | CALABRIA | 0.04 | 0.83 | density | -.093 | .882 | .200 | .747 | -0.84 | 0.09 |
|  |  |  |  | number | .984 | .002 | 1.000 | .000 | -0.85 | 0.39 |
| 3 | CAMPANIA | 1.93 | 1.82 | density | .984 | .002 | .900 | .037 | 2.22 | 2.01 |
|  |  |  |  | number | .987 | .002 | 1.000 | .000 | 1.49 | 1.59 |
| 4 | EMILIA ROMAGNA | -1.05 | -0.05 | density | .104 | .790 | .250 | .516 | 0.39 | 1.09 |
|  |  |  |  | number | .646 | .060 | .517 | .154 | 1.65 | 2.07 |
| 5 | FRIULI VENEZIA GIULIA | -0.05 | -0.33 | density | .610 | .390 | .200 | .800 | 1.37 | 1.49 |
|  |  |  |  | number | .431 | .569 | .400 | .600 | 0.45 | 0.79 |
| 6 | LAZIO | 2.27 | 2.03 | density | .978 | .004 | .900 | .037 | 1.73 | 1.76 |
|  |  |  |  | number | .999 | <.0001 | .900 | .037 | 2.21 | 2.00 |
| 7 | LIGURIA | 1.46 | 1.51 | density | .921 | .079 | .800 | .200 | 1.10 | 1.37 |
|  |  |  |  | number | .992 | .008 | .800 | .200 | 1.56 | 1.59 |
| 8 | LOMBARDY | 1.48 | 2.33 | density | .476 | .118 | .350 | .265 | 1.40 | 2.55 |
|  |  |  |  | number | .917 | <.0001 | .839 | .001 | 4.99 | 3.50 |
| 9 | MARCHE | -0.42 | 0.60 | density | -.106 | .865 | -.100 | .873 | -0.44 | 0.25 |
|  |  |  |  | number | .768 | .129 | .800 | .100 | -0.44 | 0.34 |
| 10 | PIEDMONT | 4.01 | 2.99 | density | .709 | .049 | .405 | .320 | -0.26 | 1.09 |
|  |  |  |  | number | .992 | <.0001 | .905 | .002 | 3.99 | 2.98 |
| 11 | PUGLIA | -0.48 | 0.80 | density | -.056 | .916 | .086 | .872 | 1.03 | -1.18 |
|  |  |  |  | number | .730 | .100 | .600 | .208 | 0.95 | 1.36 |
| 12 | SARDINIA | 1.76 | 1.78 | density | .021 | .953 | .800 | .104 | 2.23 | 2.01 |
|  |  |  |  | number | .796 | .107 | 1.000 | .000 | -1.06 | -1.05 |
| 13 | SICILY | 0.96 | 1.45 | density | .691 | .039 | .617 | .077 | 0.39 | 0.25 |
|  |  |  |  | number | .724 | .027 | .483 | .187 | 0.12 | 1.46 |
| 14 | TUSCANY | 4.27 | 3.18 | density | .075 | .837 | .406 | .244 | 2.97 | 2.44 |
|  |  |  |  | number | .934 | <.0001 | .552 | .098 | 4.93 | 3.42 |
| 15 | VENETO | -0.30 | -0.17 | density | .775 | .041 | .607 | .148 | -0.20 | -0.93 |
|  |  |  |  | number | .884 | .008 | .893 | .007 | -0.46 | -1.29 |

$cov$ = COVID-19 cases, $t_k$ = ratio between kurtosis and standard deviation of kurtosis, $t_s$ = ratio between skewness and standard deviation of skewness, $R$ = Pearson's correlation value, $r$ = Spearman's correlation value, $p$ = p-value.

### Speed of spread of COVID-19: Correlation with latitude and minimum temperatures

We found a significant strong correlation between the angular coefficients *b* of the various regions and their latitude (Table 2). These data indicate the dependence of COVID-19 on geographical and/or climatic factors (*R* = .895, *p* < .0001, *r* = .874, *p* < .0001). In particular, we found a significant correlation with the historical averages (last 30 years) of the minimum temperatures in the regions (*R* = −. 576, *p* = .039; *r* = −. 629, *p* = .021 for March, *R* = −. 685, *p* = .010, *r* = −. 615, *p* = .025 for February). By narrowing the analysis to the regions that showed a net correlation, we obtained much stronger correlations (*R* = −. 849, *p* = .008, *r* = −. 940, *p* = .005 for March, *R* = −. 923, *p* = .001, *r* = −. 872, *p* = .005 for February). The same is true for the correlation with latitude (*R* = .926, *p* = .001, *r* = .886, *p* = .003).

**Table 2.** Geographical and environmental data of the regions in which a correlation was found (including suspicious ones).

| **n** | **Region** | **Latitude** | ***b*** | **min T** |
| --- | --- | --- | --- | --- |
| **1** | CALABRIA | 39 | 0.00068015 | 7 |
| **2** | CAMPANIA | 40.5 | 0.00092375 | 6 |
| **3** | EMILIA ROMAGNA | 44.5 | 0.004838 | 4 |
| **4** | LAZIO | 42 | 0.0015029 | 4 |
| **5** | LIGURIA | 44.5 | 0.0072048 | 8 |
| **6** | LOMBARDIA | 45.5 | 0.0077793 | 2 |
| **7** | MARCHE | 43.5 | 0.0067591 | 4 |
| **8** | PIEMONTE | 45 | 0.0069997 | 3 |
| **9** | PUGLIA | 41 | 0.0011406 | 7 |
| **10** | SARDEGNA | 40 | 0.0019463 | 6 |
| **11** | SICILIA | 37.5 | 0.00046783 | 9 |
| **12** | TOSCANA | 43.5 | 0.0034741 | 5 |
| **13** | VENETO | 45.5 | 0.0044597 | 4 |
|  | Kurtosis t | -0.72 | -1.26 | -0.54 |
|  | Skewness t | -0.76 | 0.41 | 0.38 |

*b* = angular coefficient of the straight line that interpolates the COVID-19 cases and the number of inhabitants per region, *min* = minimum value, *T* = temperature.

### Correlation between COVID-19 cases and PM10 and PM2.5 daily averages (Lombardy only)

We identified a strong significant correlation between the number of COVID-19 cases until August 12 and the average daily concentrations of PM2.5 in Lombardy until February 29, 2020 (r=.76, p=.004). We found no significant correlation with PM10 in the same periods. Therefore, in the long run, the correlation with PM2.5 was more statistically incisive than that with PM10. However, in the early stages of the outbreak (until February 26, 2020), we found a correlation with PM2.5 (*r*= .63, *p* = .029) and PM10 (*r* = .72, *p* = .009). In the second week of March, the correlation with PM10 disappeared, while that with PM2.5 continued to exist until now. We identified a drastic surge in COVID-19 cases near 40 *μg*/*m*^3^ PM2.5 and 50*μg*/*m*^3^ PM10 (Figure 1); therefore, these may be thresholds beyond which particulate pollution clearly favors the spread of SARS-CoV-2. All the correlations found are statistically valid as they are not related to the other quantities analyzed.

**Figure 1.**
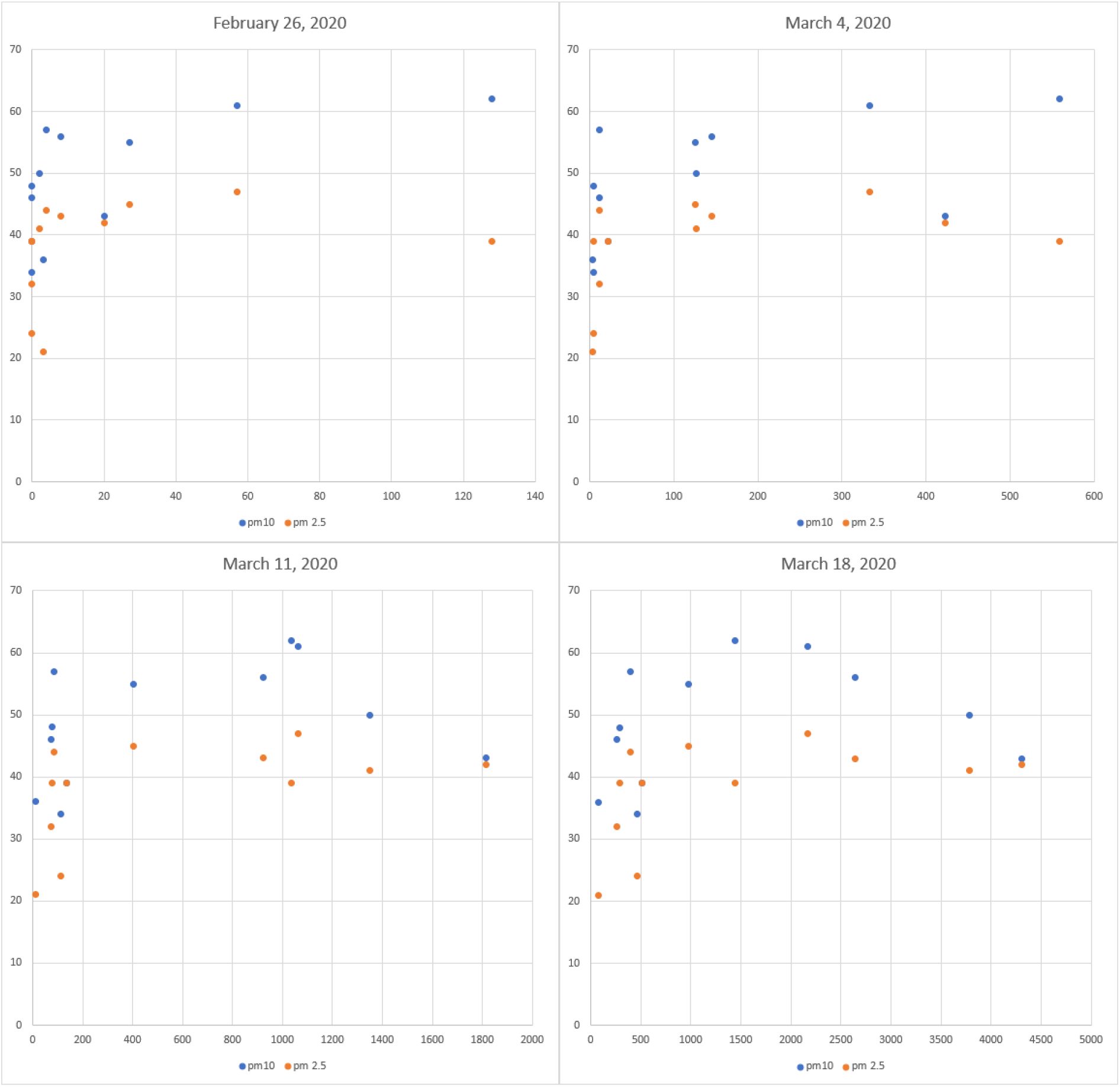
COVID-19 cases – PM10 and PM2.5 scatterplot evolution from February 26 to March 18, 2020.

### Correlation between COVID-19 cases and population density

We found significant strong correlations between COVID-19 cases and population density in % of regions such as Abruzzo (4), Campania (1), Lazio (1), Sicily (2), and Veneto (1) (*R*_*best*_ = .880, 95% *CI*: .757 – 1.000, *p*_*best*_ = .023, 95% *CI*: .006 - .040). Suspicious correlations have been found in Piedmont and Liguria (Table 1). The only pure correlation was that in Abruzzo since there was a correlation between the population density and number in all other cases (*R*_*best*_ = .939, 95% *CI*: .880 – 997, *p*_*best*_ = .003, 95% *CI*: .001 - .006). However, we highlighted discrepancies between the onset of the above in Sicily and Veneto.

### Plausible scenarios

The results are compatible with the following scenarios:

1. The almost immediate correlations between COVID-19 cases and the number of inhabitants per province in Campania, Lazio, Sicily, Liguria, and Piedmont strongly indicate that SARS-CoV-2 was in circulation for a long time before the first confirmed case. Observing the Lombardy trend, we deduced that the virus seems to have taken four weeks to correlate with the demographic dimension. It is therefore plausible that COVID-19 spread in Italy from January 2020 (or earlier).
2. Low temperatures can weaken the immune defenses, favoring the contagion from SARS-CoV-2.
3. Low temperatures can push people to have gatherings indoors and without air circulation, favoring the spread of the novel coronavirus.
4. Low temperatures could promote the survival of the virus.
5. PM2.5 can weaken the immune defenses, favoring the contagion from novel coronavirus.
6. PM2.5 can serve as an excellent carrier for the novel coronavirus.
7. High concentrations of PM10 may have contributed to the spread of the virus by acting as a carrier. This is supported not only by the initial correlation with PM10 but also by the extremely high PM10 concentration found in the first outbreak in Lodi (67 /).
8. It is possible that SARS-CoV-2 has undergone evolutionary mutations in northern Italy

(particularly in Lombardy). This agrees with the possibility that COVID-19 spread nationwide, mistaken for flu or common colds until the aforementioned mutation occurred.

## Discussion

The first two confirmed cases of COVID-19 in Italy were two Chinese tourists who landed on January 19, 2020 [5]. They could travel freely around the town, making a brief stop in Parma and staying in a hotel in Rome [11]. We believe that it is statistically unlikely and morally incorrect to associate them with the arrival of the novel coronavirus in Italy. In particular, our results suggest that SARS-CoV-2 has been circulating in Italy since early January, probably mistaken for flu or the common cold. In fact, having ascertained that:

- Those suffering from COVID-19 were largely asymptomatic [12]
- Symptoms associated with COVID-19 are milder in children compared with adults [13]
- The incubation period ranges from 2 to 14 days with an average of 5–6 days [14]
- 80% of people with COVID-19 have mild symptoms [15]
- Average mortality is about 1% [16, 17]
- The fatality rate is proportional to age and extremely high for patients over 65 years [18]

the most likely hypothesis is that the virus had been present in Italy before their arrival. All these factors drastically increase the time needed to identify and isolate a person infected with COVID-19 since: a) the asymptomatic individuals infected without their knowledge and the people with whom they have been in contact, b) children (besides the problem of asymptomaticity) showed milder symptoms than adults, inducing parents and relatives to associate them with normal colds or flu, c) the symptoms were identified up to 14 days after contagion, allowing infected subjects to infect other people unknowingly, d) in the overwhelming majority of cases, symptomatic patients showed mild symptoms not causing concern in work colleagues and relatives, and e) the fact that the death risk was extremely concentrated in the high age groups prevented an easy assessment of the extent of the epidemic since these groups were naturally more exposed to fatal phenomena, that is, until high numbers have been reached, the collective psychological impact was low. The marked correlation between the speed of the spread of the virus among the population and the minimum temperatures of each region suggests that the late autumn and winter seasons can strongly favor the pandemic. In fact, due to low external temperatures, people more frequently have indoor gatherings without recirculation of air, which creates favorable conditions for the proliferation of viruses [19]. Just as rhinoviruses, adenoviruses, and influenza viruses, the novel coronavirus may also survive better in colder and drier climates [20-23]. Furthermore, sudden changes in temperature and cold can lower immune defenses [22]. The strong and prolonged correlations we found with fine PM2.5 in the Lombardy region indicate that this type of pollution played an important role in the epidemic. This may be linked to the fact that fine particles substantially reduce immune defenses as well as increase the severity of symptoms due to damage induced in the respiratory system [24, 25]. Moreover, PM2.5 (and PM10) could act as a virus carrier [26]. In fact, from Figure 1, it can be seen that there is a drastic increase in the number of COVID-19 cases when PM2.5 and PM10 exceed approximately 40 *μg*/*m*^3^ and 50 *μg*/*m*^3^, respectively. These could be the thresholds beyond which particulate matter significantly favors the spread of SARS-CoV-2. In addition to empirical data and other studies, this hypothesis is consistent with the first outbreak in Lodi, where the PM10 average daily concentration was the highest of the month (67 *μg*/*m*^3^) [7, 27]. However, our results show a greater incisiveness of PM2.5 compared to PM10 in the spread of the virus in Lombardy (Figure 1). Finally, considering that in Lombardy, the correlation between COVID-19 cases and the number of inhabitants per region became significant after four weeks, the severity of symptoms was more severe than in other regions, and the basic reproduction number (*R*0) seems to have been the highest, we suggest that an evolutionary genetic mutation may have occurred in Lombardy [7]. In fact, although the genome does not appear to have changed substantially, Zhang et al. showed that even small mutations can cause significant changes in SARS-CoV-2 behavior [28].

### Limitations

Statistical correlations can provide valid support for hypotheses and theories as well as fundamental indicators of phenomena to be explored; however, they cannot demonstrate the causal nature of a phenomenon.

## Conclusion

We found significant strong and lasting correlations between the spread of SARS-CoV-2 and the number of inhabitants of each region, between the spread-speed of COVID-19 and the historical minimum temperatures in the months of February and March, and between the number of COVID-19 cases and the average daily concentrations of fine PM2.5. Correlations with the average daily concentrations of PM10 were found up to the first week of March, indicating that this type of pollution also played a role in the spread of the virus, linked to exceeding specific daily peaks. Therefore, we suggest that the health authorities pay particular attention to the arrival of the winter months, not only by investing in adequate precautions but also by launching various awareness campaigns on air recirculation indoors and in avoiding exposure to the cold. In addition, it will also be necessary to carefully monitor levels of PM10 and 2.5, even by limiting the use of cars if necessary.

## Data Availability

All the data necessary for carrying out this study are presented in the paper or in the articles reported in the references.

